# Transition from non-diabetes to diabetes: a population-based longitudinal study in Vietnamese individuals

**DOI:** 10.1101/2025.09.15.25335785

**Authors:** Lan T. Ho-Pham, Linh D. Mai, Thach S. Tran, Tuan V. Nguyen

**Author notes:** **Corresponding Authors** Lan T. Ho-Pham, Tuan V. Nguyen.

## Abstract

**Objective:** This study sought to determine the incidence of T2DM and identify risk factors among a Vietnamese urban cohort without previous T2DM diagnoses to support early intervention.

**Methods:** This population-based prospective study included 1971 individuals (1368 women) free of T2DM at baseline. Participants were re-assessed after a median follow-up of 2.0 years. Glycosylated hemoglobin (HbA1c) levels were measured using high-pressure liquid chromatography (HPLC) analyzers (ADAMS A1c HA-8160, Arkray, Kyoto, Japan). T2DM was defined as an HbA1c value ≥ 6.5%, while prediabetes was defined as an HbA1c value between 5.7% and 6.4%. Incident T2DM was identified as newly developed diabetes from a non-diabetic status. The association between potential risk factors and T2DM was analyzed using logistic regression.

**Results:** At baseline, 1137 participants (58%) were classified as ‘Normal’ and 834 (42%) as ‘Pre-diabetes’. Over the follow-up, 16% of initially normal individuals progressed to prediabetes, and 10% of those with prediabetes progressed to diabetes. Overall, 4.3% transitioned to diabetes. Advancing age (OR per year: 1.03; 95% CI, 1.01 to 1.06) and higher BMI (OR per unit: 1.20; 95% CI, 1.12 to 1.27) were significantly associated with an increased risk of developing T2DM. A predictive model incorporating age, body mass index (BMI), and baseline HbA1c resulted in an area under the ROC curve of 0.90.

**Conclusions:** The study revealed a notable 4.3% incidence of T2DM over two years among Vietnamese adults. Significant predictors included age and BMI, underscoring the need for targeted preventive measures focusing on weight management and monitoring in high-risk groups.

## Introduction

Type 2 diabetes mellitus (T2DM) has undergone a dramatic rise in Vietnam, transitioning from a relatively uncommon condition to a major public health concern. Population-based studies observed a rapid doubling of prevalence from 2.7% in 2002 to 5.4% in 2012 [1], mirroring a global trend. In our previous study conducted in Ho Chi Minh City, we found that the prevalence of T2DM and pre-diabetes was 12% and 40%, respectively [2], which is comparable to levels observed in the United States [3]. This rapid increase imposes a substantial burden on healthcare systems, as T2DM complications, such as cardiovascular disease, kidney failure, and blindness, lead to significant morbidity and mortality [4].

Understanding the dynamics of this disease, particularly its incidence of new cases in the population, is crucial for developing effective preventive strategies. While numerous studies worldwide have explored T2DM incidence, the focus has often been on economically developed nations [5]. In the Framingham Heart Study, the aged-adjusted annual incidence of T2DM was about 4% in women and ∼6% in men during the 1990s [6]. In adolescents without T2DM but with overweight and obesity, the incidence of T2DM was much lower: 2.1 per 1000 person-years [7]. Data from Asia, particularly prospective studies that follow individuals over time to identify new cases, remains scarce. This lack of region-specific data hinders the development of targeted interventions that address the unique risk factors and demographics of the Vietnamese population.

This study sought to fill this knowledge gap by estimating the incidence of both pre-diabetes and T2DM in Vietnam. We employed a robust prospective cohort design, meticulously following a population-based sample of Vietnamese adults over a defined period. Our aims were two-fold: first, we wanted estimate the incidence rates of pre-diabetes and T2DM, providing valuable insights into the disease trajectory within the Vietnamese population; and second, we leveraged the data to develop a robust prediction model to identify individuals at high risk of developing T2DM, allowing for early detection and preventative measures.

## Study Design and Methods

### Study Design

This study is part of the Vietnam Osteoporosis Study (VOS) project, with its design and procedures previously published [8]. Briefly, VOS is a prospective, population-based investigation that commenced in 2015 in Ho Chi Minh City, Vietnam, and has followed participants biennially through 2024. The study’s procedures and protocols were approved by the research and ethics committee of the People’s Hospital 115 on August 6, 2015 (approval number: 297/BV-NCKH). Written informed consent was obtained from all participants.

We employed two approaches to recruit participants. In the first approach, we collaborated with community organizations to obtain a list of members. From this list, a computer program randomly selected individuals who met the age and sex criteria. Selected individuals were then sent a letter inviting them and their family members to participate in the study. The second approach involved recruiting participants through television, the Internet, and flyers in universities. The flyers, written in Vietnamese, outlined the study’s purposes, procedures, and benefits for participants. Those who agreed to participate were transported to the Bone and Muscle Research Laboratory at Ton Duc Thang University for clinical assessment and evaluation. Participants did not receive any financial incentive but were provided with a free health check-up and lipid analyses.

For the present analysis, we included participants aged 30 years and above who had blood samples collected both at baseline and at a follow-up visit approximately two years later (between December 2015 and January 2020). We (the principal authors) had access to identifiable participant information during and after data collection. Individuals with a history of diabetes or those who had used diabetic drugs were excluded. Additionally, we excluded participants who were classified diabetes by the American Diabetes Association (ADA) criteria for diabetes [9].

### Data Collection

We used a structured questionnaire to collect extensive data on demographic factors, medical history, lifestyle factors, physical activity, and dietary habits at baseline and each subsequent visit. For lifestyle factors, smoking was classified into past smoking and current smoking, whereas alcohol intake was quantified as average numbers of standard drinks per day at present and within the last 5 years. Anthropometric factors, including weight, height and waist circumference, were also measured using the standard protocol [8]. Body mass index was calculated from the weight in kilograms divided by the square of the height in meters. The participants were classified into 4 groups: underweight if BMI <18.5 kg/m^2^, normal if BMI between 18.5 and 22.9 kg/m^2^, overweight if BMI between 23 and 24.9 kg/m^2^, and obese if BMI >25 kg/m^2^.

Overnight fasting venous blood samples were collected from each participant at baseline and the subsequent visits. The serum was immediately frozen to −20°C, and the biochemical analysis was taken within 24 hours after the collection. Glycosylated hemoglobin (HbA1c) levels were quantified using high pressure liquid chromatography (HPLC) analyzers ADAMS A1c HA-8160 (Arkray, Kyoto, Japan). The intra- and interassay coefficient of variation for this method is less than 1%. Fasting plasma glucose (FPG) was measured by the hexokinase method (Advia 1800 Autoanalyzer; Bayer Diagnostics, Leverkusen, Germany) with an intra-measurement coefficient of variance of 0.98%-1.34%.

Pre-diabetes and diabetes mellitus were ascertained using the American Diabetes Association 2023 Criteria [9]. Specifically, diabetes mellitus was defined as HbA_1c_ value ≥ 6.5% (48 mmol/mol) or FPG ≥ 126 mg/dL (7 mmol/L), whereas a participant was diagnosed to have prediabetes if his/her HbA1c value between 5.7% and 6.4% (39-46 mmol/mol) or FPG between 100 and 125 mg/dL (5.6 - 6.9 mmol/L). Incident diabetes mellitus was defined as any diabetes mellitus newly developed from non-diabetic status, including euglycemia or prediabetes at baseline during the follow-up.

### Data Analysis

The incidence of prediabetes and diabetes was determined by counting the cases of new-onset prediabetes and diabetes that occurred after the baseline over the follow-up period. The corresponding confidence interval was estimated using the binomial assumption. Incidence rates were also stratified by sex and age groups (40-49, 50-59, 60-69, and 70+ years).

In the subsequent analysis, we investigated which baseline factors were associated with the incidence of T2DM. We employed a multiple logistic regression model, treating new-onset T2DM as the outcome and considering the following variables as potential predictors: age, gender, baseline HbA1c, blood glucose levels, BMI, WHR, smoking status, and alcohol consumption. Given the presence of multiple potential predictors and numerous possible models, we used the Bayesian Model Averaging (BMA) approach to identify the optimal model. The BMA approach is statistically more robust than a conventional stepwise approach for identifying an optimal prediction model. We evaluated the model’s discrimination performance using Harrell’s C index, and its corresponding 95% CI was reported. In the next analysis, we aimed to determine the baseline HbA1c value that indicates a high risk of progressing to T2DM. To achieve this, we employed change-point regression analysis [10]. All statistical analyses were conducted using the R Statistical Environment [11]. The datasets generated during and/or analyzed in the current study are available from the corresponding author upon reasonable request.

## Results

Ultimately, the analysis included 1368 women and 603, all aged 30 years and older at baseline and without T2DM (**Table 1**). As expected, men were on average taller and heavier than women, with statistically significant differences in height (163 cm vs. 153 cm) and weight (62.0 kg vs. 53.5 kg). Women had a higher percentage of body fat (42.3% vs. 30.6%) and lower lean mass (31.1 kg vs. 43.1 kg) compared to men. Interestingly, the average HbA1c levels were similar between genders. Health behavior differences were marked, with a significantly higher percentage of men being current smokers (46.1% vs. 1.0%) and alcohol users (45.9% vs. 2.9%).

**Table 1.** Characteristics of 1368 women and 603 men aged 30 years and older at baseline.

| Variable | Women | Men | P-value |
| --- | --- | --- | --- |
| Number of subjects | 1368 | 603 |  |
| Age (years) | 53.7 (9.3) | 53.8 (9.7) | 0.864 |
| Height (cm) | 153 (5.2) | 163 (6.1) | <0.0001 |
| Weight (kg) | 53.5 (8.0) | 62.0 (9.7) | <0.0001 |
| BMI (kg/m<sup>2</sup>) | 23.0 (3.1) | 23.3 (3.2) | 0.023 |
| BMI classification |  |  | 0.033 |
| • Underweight | 64 (4.7) | 31 (5.1) |  |
| • Normal | 999 (73.0) | 397 (65.8) |  |
| • Overweight | 267 (19.5) | 159 (26.4) |  |
| • Obese | 37 (2.7) | 16 (2.7) |  |
| Percent body fat (%) | 42.3 (4.4) | 30.6 (5.3) | <0.0001 |
| Fat mass (kg) | 23.1 (5.2) | 19.4 (5.6) | <0.0001 |
| Lean mass (kg) | 31.1 (3.9) | 43.1 (5.3) | <0.0001 |
| HbA<sub>1c</sub> (%) | 5.59 (0.35) | 5.59 (0.34) |  |
| HbA<sub>1c</sub> classification |  |  |  |
| • Normal | 791 (57.8) | 346 (57.4) | 0.894 |
| • Pre-diabetes | 577 (42.2) | 257 (42.6) |  |
| Current smoking (%) | 13 (1.0) | 278 (46.1) | <0.0001 |
| Current use of alcohol (%) | 39 (2.9) | 277 (45.9) | <0.0001 |

**Table 2** shows how individuals transitioned between normal, pre-diabetes, and diabetes states over a two-year period. Among those initially classified as normal, 83.9% remained normal, 16.0% developed pre-diabetes, and 0.1% progressed to T2DM. For those with pre-diabetes at baseline, 32.0% reverted to normal, 58.0% remained pre-diabetic, and 10.0% developed T2DM. Overall, combining normal and pre-diabetes groups, 4.3% of individuals developed T2DM within the follow-up period.

**Table 2.**
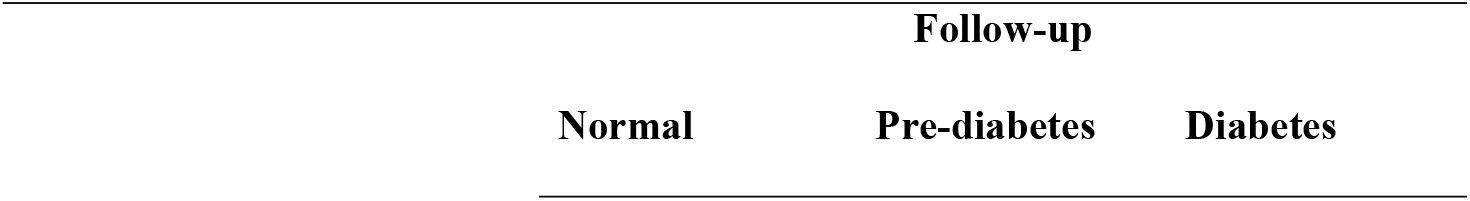
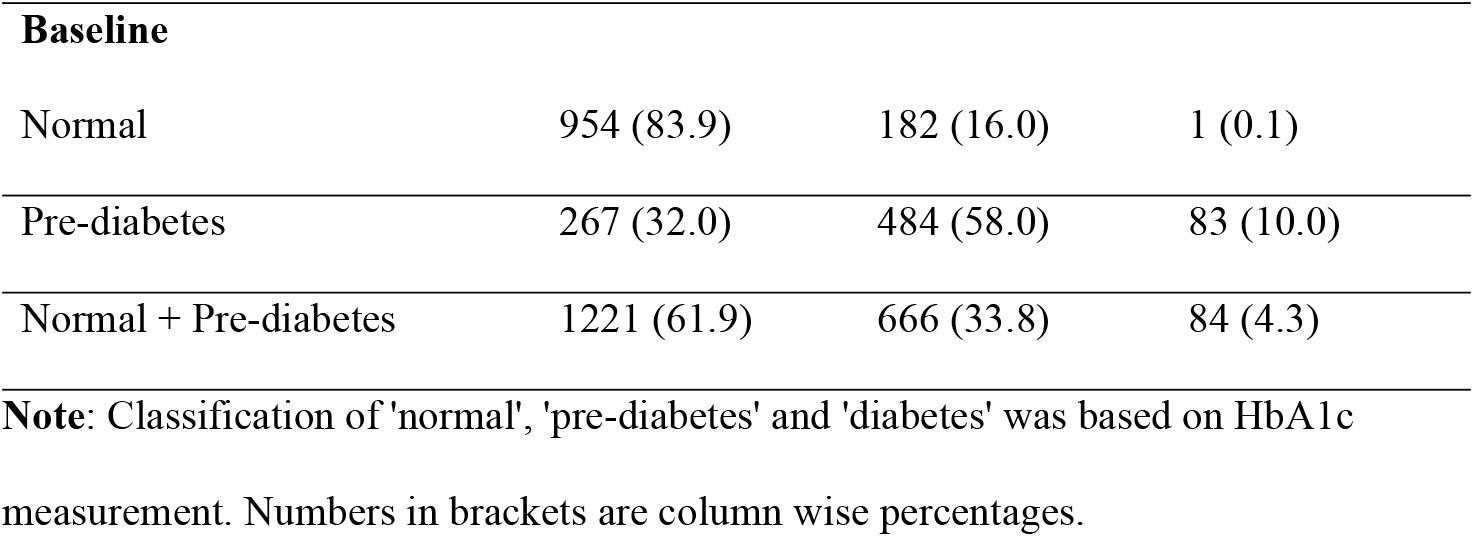
The transition between baseline and follow-up diabetic states.

|  | Follow-up |  |  |
| --- | --- | --- | --- |
|  | Normal | Pre-diabetes | Diabetes |

| **Baseline** |  |  |  |
| --- | --- | --- | --- |
| Normal | 954 (83.9) | 182 (16.0) | 1 (0.1) |
| Pre-diabetes | 267 (32.0) | 484 (58.0) | 83 (10.0) |
| Normal + Pre-diabetes | 1221 (61.9) | 666 (33.8) | 84 (4.3) |

**Note:** Classification of 'normal', 'pre-diabetes' and 'diabetes' was based on HbA1c measurement. Numbers in brackets are column wise percentages.

The incidence of T2DM increased with advancing age (**Table 3**). For instance, in the 30-49 age group, 2.3% developed T2DM, whereas in those aged 50 years and older, 5.3% progressed to T2DM.

**Table 3.** Incidence of type 2 diabetes by age group.

|  | **Follow-up (2 years)** |  |  |
| --- | --- | --- | --- |
|  | **Normal** | **Pre-diabetes** | **Diabetes** |
| 30 - 39 | 76 (80.0) | 16 (16.8) | 3 (3.2) |
| 40 - 49 | 446 (74.7) | 138 (23.1) | 13 (2.2) |
| 50 - 59 | 463 (57.4) | 304 (37.7) | 40 (5.0) |
| 60 - 69 | 183 (52.7) | 142 (40.9) | 22 (6.3) |
| 70+ | 53 (42.4) | 66 (52.8) | 6 (4.8) |
| All | 1221 (61.9) | 666 (33.8) | 84 (4.3) |

**Note:** Classification of 'normal', 'pre-diabetes' and 'diabetes' was based on HbA1c measurement. Numbers in brackets are columnwise percentages.

The correlation between baseline and follow-up HbA1c levels was moderate, with a correlation coefficient of 0.65 (**Figure 1**). To identify a baseline HbA1c threshold at which the risk changes, we applied a change-point regression model to the data. The analysis indicated that the threshold was 5.6 (95% CI, 5.5 to 6.2).

**Figure 1:**
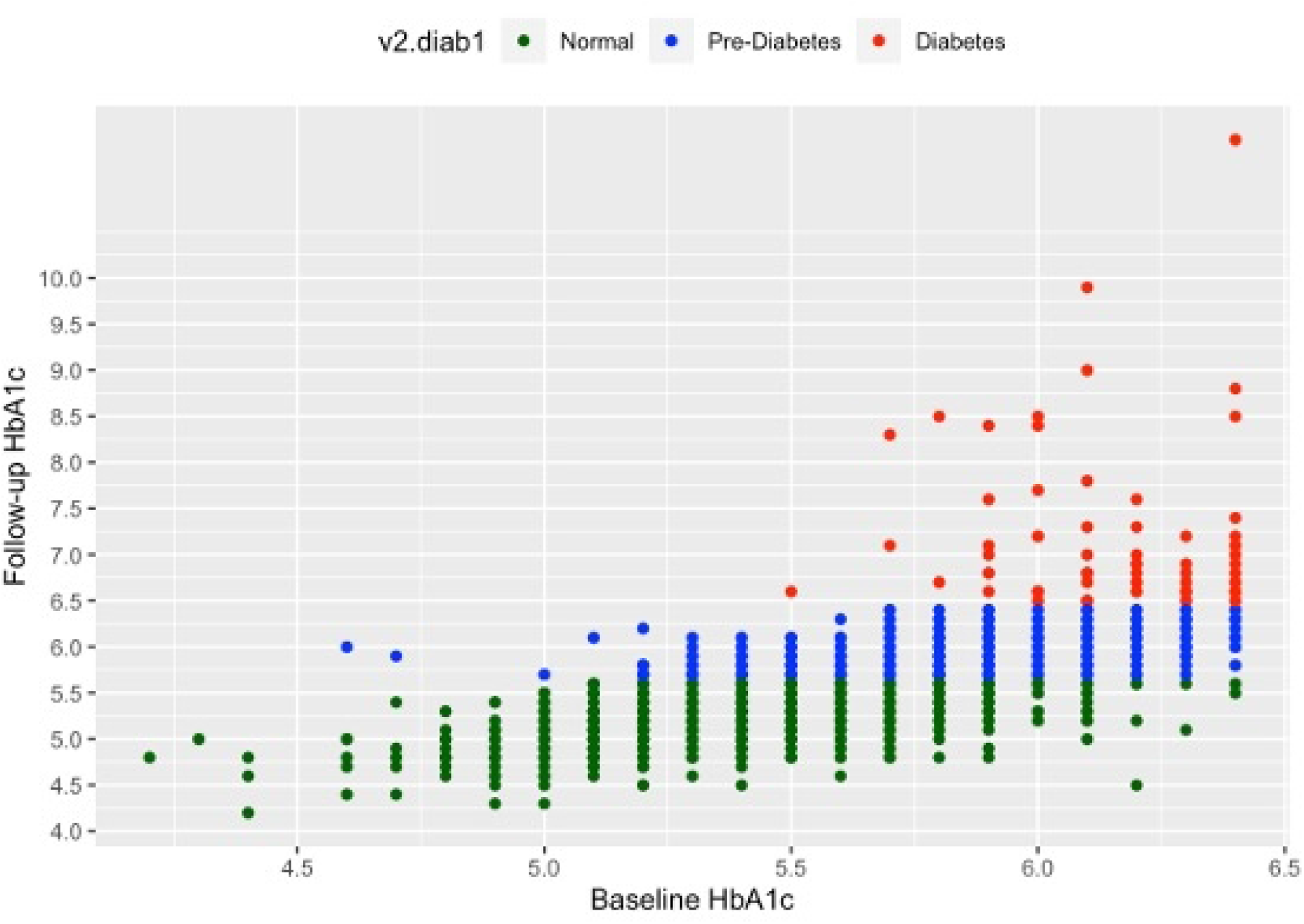
The relationship between baseline HbA1c and follow-up HbA1c stratified by diabetic status. Green dots represent individuals who maintained a normal HbA1c level, blue dots denote those in the pre-diabetes range, and red dots indicate individuals who have transitioned to or remained in the diabetes range.

**Table 4** presents the parameters of potential models for predicting the risk of developing T2DM. Model 1 incorporated age and BMI, while Model 2 included baseline HbA1c and BMI. Each additional year of age was associated with a 3% (95% CI, 1% to 6%) increase in the odds of developing T2DM. Additionally, each unit increase in BMI correlated with an approximately 20% (95% CI, 12% to 27%) rise in the odds of T2DM. In Model 2, a 0.1% increase in HbA1c was linked to nearly a twofold increase in the odds of diabetes. Regarding the area under the ROC curve (AUC), Model 2 (AUC = 0.94) significantly outperformed Model 1 (AUC = 0.69).

**Table 4.** Risk factors for predicting the risk of progression to type 2 diabetes.

| Risk factor | Unit | Odds ratio (95% CI) | P-value |
| --- | --- | --- | --- |
| **Model 1** |  |  |  |
| Age | +1 yr | 1.03 (1.01 - 1.06) | 0.004 |
| BMI | + 1 kg/m<sup>2</sup> | 1.19 (1.12 - 1.27) | < 0.001 |
| **Model 2** |  |  |  |
| Baseline HbA1c | +0.10 | 1.92 (1.73 - 2.14) | < 0.001 |
| BMI | + 1 kg/m<sup>2</sup> | 1.13 (1.05 - 1.22) | < 0.001 |

The area under the ROC curve was 0.69 for model 1 and 0.94 for model 2.

## Discussion

Despite the growing prevalence of T2DM as a significant public health issue in the Asian populations, few prospective studies have investigated the incidence of T2DM over time. To address this knowledge gap, we conducted a longitudinal study in Vietnam, and found that among individuals without a prior diagnosis of T2DM, the incidence of developing T2DM over a two-year period was 4.3%. Additionally, advancing age and high BMI were independently associated with a higher risk of developing T2DM. These findings deserve further elaboration.

Our results align with previous research that identifies age and BMI as critical risk factors for type 2 diabetes. For instance, a study by Hu et al. [12] demonstrated a similar trend, indicating that higher BMI and older age were strong predictors of diabetes incidence. However, our observed incidence rate of 4.3% over two years is somewhat lower than the 5.9% reported in the Framingham Offspring Study [6], which could be attributed to differences in population demographics and study design. Furthermore, a meta-analysis [13] reported a wide range of incidence rates across different populations, further suggesting that regional, ethnic, and lifestyle variations significantly influence diabetes risk.

The discrepancies between our findings and those of other studies could be due to several factors. Firstly, our study population may differ in terms of genetic predispositions, lifestyle factors, and baseline health status. Our cohort consisted primarily of individuals from urban settings with access to healthcare, which may not be representative of rural populations or those with limited healthcare access. Additionally, variations in diagnostic criteria and methods of data collection can lead to differences in reported incidence rates. Our study utilized stringent diagnostic criteria and a robust follow-up protocol, which may have contributed to a more accurate but slightly lower incidence rate. Moreover, the temporal scope of our study, focusing on a recent two-year period, might reflect improvements in public health interventions and awareness campaigns that have mitigated diabetes risk factors to some extent.

The link between BMI and type 2 diabetes is well-documented and can be explained by several biological mechanisms. Higher BMI is often associated with increased adiposity, which leads to insulin resistance—a key pathophysiological feature of type 2 diabetes. Adipose tissue, particularly visceral fat, secretes various adipokines and pro-inflammatory cytokines that impair insulin signaling and glucose metabolism [14]. Additionally, obesity is linked to a chronic low-grade inflammatory state, which exacerbates insulin resistance. The accumulation of ectopic fat in the liver and muscles further disrupts insulin action and glucose homeostasis. Moreover, increased BMI is often correlated with other metabolic disturbances, such as dyslipidemia and hypertension, which further elevate diabetes risk. These interconnected pathways highlight the complex interplay between obesity and metabolic health.

The continuous association between HbA1c levels and the risk of T2D is well-established. However, determining the specific HbA1c threshold that indicates a ‘high risk’ of developing T2DM remains a critical question. Our change-point regression analysis suggested that this threshold is 5.6%. This finding aligns closely with the ADA’s recommendation [15] and corroborates previous study [16].

The findings of this study have significant public health implications. Given the strong association between age, BMI, and diabetes risk, targeted interventions aimed at weight management and lifestyle modifications in older adults could substantially reduce the incidence of type 2 diabetes. Health care providers should prioritize regular screening and preventive measures for individuals at higher risk, particularly those with elevated BMI. Early intervention strategies, including dietary counseling, physical activity promotion, and behavioral therapy, can be effective in mitigating these risk factors. Additionally, our study highlights the need for public health policies that promote healthy aging and weight control as critical strategies in combating the diabetes epidemic. Community-based programs that support healthy eating and active living can play a vital role in reducing the burden of diabetes. Furthermore, our findings suggest that personalized healthcare approaches, considering individual risk profiles, can enhance the effectiveness of prevention and management strategies.

Our study has several strengths that contribute to the robustness and reliability of the findings. Firstly, the large sample size enhanced the statistical power of the study, allowing for more precise estimates of the incidence of type 2 diabetes. Additionally, participants were drawn from the community rather than hospitals, which helped avoiding selection bias that might occur if the sample was limited to those already seeking medical care. Furthermore, the measurements of HbA1c were conducted using state-of-the-art technology, ensuring high accuracy and reliability of the data collected. However, the study is not without its limitations. The participants were predominantly from an urban setting, which may not be representative of the rural population. This urban bias could limit the generalizability of the findings to rural areas, where lifestyle and health determinants might differ significantly. Therefore, future studies should aim to include a more diverse sample that encompasses both urban and rural populations to provide a more comprehensive understanding of the incidence and risk factors of type 2 diabetes across different settings.

## Conclusion

In conclusion, our study demonstrates a 4.3% incidence of diabetes over a two-year period among individuals without a prior diagnosis, with increased age and BMI being significant risk factors. The strong association between BMI and diabetes underscores the critical role of weight management in diabetes prevention. Public health initiatives and clinical interventions targeting weight control and healthy aging could substantially reduce the burden of type 2 diabetes.

## Data Availability

Some or all datasets generated during and/or analyzed during the current study are not publicly available but are available from the corresponding author on reasonable request.

## ACKNOWLEDGMENTS

This research is funded by Foundation for Science and Technology Development of Ton Duc Thang University (FOSTECT, http://fostect.tdt.edu.vn), Grant number FOSTECT.2014.BR.09, and a grant from the Department of Science and Technology of Ho Chi Minh City. We sincerely thank Ms Tran Thi Ngoc Trang and Fr Pham Ba Lam for coordinating the recruitment of participants. We also thank doctors and medical students of the Pham Ngoc Thach University of Medicine for the data collection and clinical measurements.

